# Rapid Discrimination of High-Grade Prostate Cancer Using Label-Free Fluorescence Lifetime Measurements

**DOI:** 10.1101/2025.05.23.25328257

**Authors:** Julien Bec, Xiangnan Zhou, Yash Tipirneni, Shuai Chen, Jinyi Qi, Kenneth A. Iczkowski, Marc Dall’Era, Laura Marcu

## Abstract

**Purpose:** Histologic evaluation of prostatic needle biopsies is essential for prostate cancer (PCa) diagnosis and treatment planning, yet tissue targeting remains suboptimal despite MRI-guided Bx procedures. This pilot study investigates the use of label-free Fluorescence Lifetime Imaging (FLIm) for real-time biopsy guidance. Using ex vivo specimens, we assess FLIm’s preliminary efficacy in discriminating malignant from benign prostate tissue.

**Materials and Methods:** Twenty patients undergoing prostate biopsy were enrolled. FLIm measurements were performed immediately after sample collection using a custom fiber-optic probe. Optical parameters from 4 spectral bands associated with distinct endogenous fluorophores including structural proteins and metabolic cofactors (e.g. NADH, FAD) were extracted and labeled based on histological annotation. Data were analyzed to characterize tissue-type differences and train and evaluate a classifier to distinguish malignancy.

**Results:** Separation between benign tissue and Gleason grade ≥4 PCa was achieved using just 2 of 56 FLIm-derived parameters. A Support Vector Machine classifier using all parameters achieved a ROC of 0.88 in identifying grade 4 PCa. A reduced lifetime value in the NADH-associate band, likely due to increased free NADH from upregulated glycolysis, supports the biochemical basis for optical differentiation.

**Conclusions:** FLIm shows significant potential for high-grade PCa identification. The single-fiber approach requires minimal modification for integration into current biopsy tools, supporting its feasibility for clinical translation.

## INTRODUCTION

### Prostate Cancer (Pca) Is One of the Most Common Cancers Affecting Men

PCa is the second leading cause of cancer death in men worldwide^1^. In 2018 alone, over 1.2 million new cases of PCa were diagnosed globally, with approximately 359,000 deaths. PCa screening typically involves a prostate-specific antigen (PSA) blood test and a digital rectal exam. If either of these is abnormal, additional testing such as another biomarker or multiparametric MRI (mpMRI) for further risk stratification may be used to decide if a prostate biopsy is recommended.^23^

### Role Of and Limitations of Prostatic Biopsy (Bx)

Accurate determination of PCa stage and Gleason grade is essential for treatment planning. Despite performing about 1 million prostate biopsies annually in the US ^2^., diagnostic efficacy remains limited, with standard 12-core biopsies having up to 20% false negative rates ^3^, and 40% risk of Gleason undergrading^4^ and 90% of biopsy cores are benign. While pre-biopsy MRI improves yield, targeting remains challenging^5,6^. MRI-ultrasound fusion technologies require additional equipment ^7,8^ without demonstrating significant benefits over cognitive fusion methods ^9^. These limitations persist even in MRI-guided biopsies^10^, suggesting intratumor heterogeneity^11^ significantly impacts accuracy^6^. The main factors limiting sensitivity include small sample size relative to total prostate volume, the multifocal nature of PCa ^12^, and sparse sampling, which can lead to underestimating tumor grade.

### Label-Free Optical Imaging Technology for Prostate Biopsy Guidance

Label-free optical technologies show promise for prostate biopsy guidance by providing real-time tissue assessment without contrast agents. They can detect biochemical (fluorescence, light reflectance spectroscopy) ^13,14^, structural (optical coherence tomography^15,16^) or cellular architecture (confocal endomicroscopy^17^) differences between benign and malignant tissues and have the potential to be implemented via fiber optic to enable tissue identification at the biopsy needle tip. A major challenge, however, is achieving reliable tissue discrimination performance while maintaining the size and sampling efficiency of conventional biopsy devices.

### Fluorescence Lifetime Imaging (FLIm) for PCa detection and characterization

Label-free FLIm offers a promising approach for cancer detection by capturing changes in extracellular matrix composition ^18,19^ and cell metabolism ^20–22^, that occur during malignant transformation. These changes lead to variations in optical signature across multiple wavelengths, enabling a comprehensive characterization of neoplastic tissue, potentially enhancing diagnostic accuracy and biological understanding of PCa progression.

In this pilot study involving freshly biopsied prostate specimens from 20 patients, we investigated whether biochemical and metabolic changes among tissue types (i.e. cancer, dysplasia, healthy) produce distinct autofluorescence signatures detectable by FLIm. We developed both binary and multi-class classifiers to assess the preliminary efficacy of FLIm in identifying PCa tissue. This work is designed to assess whether FLIm provides the ability to perform real-time PCa detection, with potential to guide biopsy procedures.

## MATERIALS AND METHODS

### Study Population and Design

This study, approved by the UC Davis Institutional Review Board, was performed between May and August 2024 and included 20 patients undergoing transperineal prostate Bx. Eligible patients were offered to enroll in this study, based on FLIm instrumentation availability. No additional biopsies were collected specifically for this study.

### Pulse-Sampling FLIm System

Figure 1 illustrates the custom-built, fiber-based, point-scanning FLIm system that consists of a console and reusable fiber optic probe. In-depth characterizations of this instrument and specifications are detailed extensively elsewhere.^23–27^ Briefly, the fiber optic probe consists of a 365 µm core multimode optical fiber terminated with a 2.5-mm ball lens, leading to an expected lateral resolution of 400 µm. It is used to deliver 355 nm pulsed UV laser excitation light to biological tissue (460 Hz repetition rate). The same optical fiber is used to relay autofluorescence point measurements from the tissue regions evaluated to the FLIm system. The fiber’s proximal collection end is coupled to a wavelength selection module (WSM) which features a set of four dichroic mirrors and bandpass filters selected to capture emissions of key biological fluorophores(i.e., 390±20 nm: collagen, proteoglycans, 470±14 nm: NAD(P)H, 542±25 nm: FAD, and 629±26.5 nm: porphyrins)^20^. The optical signal from each spectral band is detected by an avalanche photodiode detector and digitized (2.5 GS/s). This pulse sampling FLIm approach is suitable for use in the presence of external illumination such as operating room lights ^26,27^.

**Figure 1.**
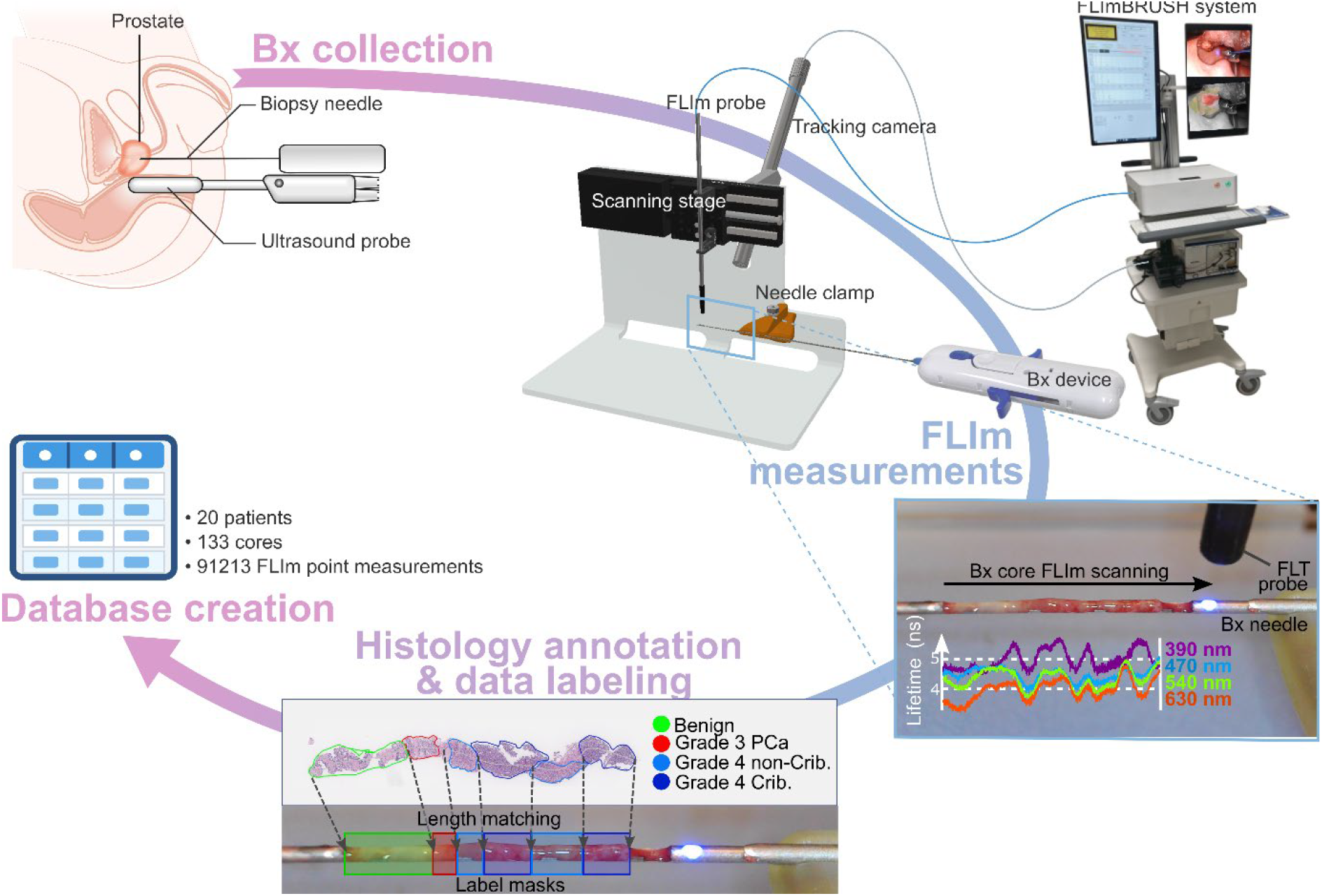
Study synopsis: Transperineal prostate biopsy collection. FLIm measurements were performed along the length of the core (2.5 mm/s) using a custom automated sample scanner to ensure consistent, high-SNR measurements over the entire length of the biopsy core. Histology slide annotation and data labeling. Creation of a clinical database matching FLIm signature and tissue type.

### Prostate Biopsy Sample Collection and Fluorescence Lifetime Data Acquisition

Once tissue was collected, the external cannula was retracted to expose the specimen, the needle of the biopsy device was positioned and secured on the scanning fixture (**Fig. 1**), and FLIm measurements were performed, all within seconds of specimen collection. A bidirectional scan provided repeat measurements of the core to assess FLIm measurement variability and evaluate potential changes of FLIm signature within seconds of specimen collection.

### FLIm Data Processing

The FLIm waveforms which originate from each spectral band were averaged four times before further processing resulting in point measurements every 22 µm. The fluorescence decay parameters from each FLIm point measurement were retrieved using a constrained least-square deconvolution with Laguerre expansion method^28^. This approach provides average lifetime values for each spectral band and captures the complex decay dynamics. For each of the four spectral bands of the instrument, the average lifetime value, spectral intensity, and 12 fitting coefficients of the Laguerre expansion were computed (Figure 2C). This leads to a total of 52 parameters (4 time-resolved average lifetimes, and 48 Laguerre expansion metrics).

**Figure 2.**
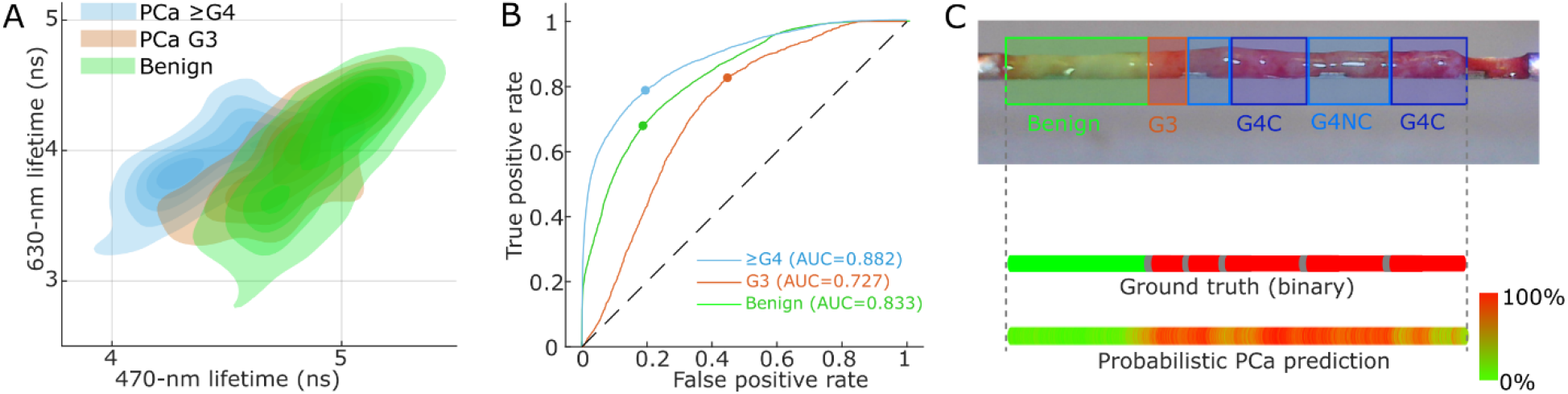
Multivariate and classification results. Contour line plot of average lifetime signature for 470-nm and 560-nm bands, for benign tissue, PCa grade 3, and PCa grade ≥4. ROC curves for 3-class SVM predictor results using 4 average lifetimes. Illustration of 2-class (PCa, benign) probabilistic classifier output demonstrating good agreement with the original labels.

### Histopathologic Processing and Annotation

Following FLIm interrogation, Bx specimens were placed on coreCARE (Uro-1, Greensboro, NC) specimen retrieval substrates, which are designed to preserve tissue integrity and orientation during the biopsy process. The samples were formalin fixed, paraffin embedded sectioned and stained (hematoxylin and eosin). High-resolution digital images of the stained cores were captured using the Aperio platform and annotated by a subspecialty-trained urologic pathologist, blinded to the FLIm data (Table 1 and Figure S1).

**Table 1.** Participants’ Clinicopathologic Characteristics.

| Variable | Result |
| --- | --- |
| Number of Patients | 20 |
| Total Cores | 133 |
| Age | 68.5+/-7.9 |
| ISUP Grade Group |  |
| GG0 | N=5 |
| GG1 | N=4 |
| GG2 | N=5 |
| GG3 | N=4 |
| GG4 | N=1 |
| GG5 | N=1 |
| Tumor grade distribution (number of FLIm point measurements/ number of patients in which it is present) |  |
| Benign (prostatic tissue, fibromuscular stroma, seminal vesicle, atrophic benign gland, high-grade prostatic intraepithelial neoplasia) | n=80614/ N=20 |
| Grade 3 | n=3936/ N=12 |
| Grade 4, not cribriform | n=4562/ N=5 |
| Grade 4 with cribriform | n=2012/ N=4 |
| Grade 5 | n=89/ N=1 |

### Database Creation

FLIm data were labeled by co-registering the FLIm measurement position along the fresh core with the annotated histologic sections. Differences in length between fresh cores and slides of the core, attributed mainly to stretching and shrinking during fixation and processing were addressed by assuming a uniform deformation of the specimen. The complete study database consisted of 91213 FLIm points obtained from 133 cores from 20 patients and included labels as well as patient level demographic and diagnostic information.

### Data Analysis and Machine Learning

Measurement repeatability was evaluated across the entire dataset by systematically evaluating differences in average lifetime for each spectral band between repeat measurements. Differences between consecutive measurements of the same specimen may originate from variability in FLIm measurements as well as potential changes in the fluorescence signature at the sample level. Variation in signal with frequency higher than the spatial resolution of the system can be attributed to measurement noise, thus an 18-point moving average was applied to lifetime traces of each repeat scan (Supplemental Fig. S2). The measurement noise intensity was computed as the standard deviation of the difference between original and smoothed traces, and the measurement noise in the smoothed traces was estimated by dividing the original noise by the square root of the number of points used for averaging. The mean absolute error (MAE) between smoothed repeat scans was calculated and compared to three times the estimated noise standard deviation in the smoothed traces. When the MAE exceeded this threshold, the difference between repeat measurements was considered statistically significant with a 99% confidence interval, indicating a genuine change in FLIm signature rather than measurement variability. To assess a potential effect on tissue classification performance, we compared the MAE between repeat scans to the standard deviation of the original measurement noise.

Multivariate analysis was initially applied to qualitatively evaluate whether FLIm-derived parameters could distinguish between tissue types. This preliminary examination focused on identifying if one or more parameters showed potential differentiation capabilities, serving as a first-pass screening before more rigorous statistical validation. Visual inspection of parameter distributions suggested that even a minimal subset of FLIm features might effectively separate tissue categories, guiding subsequent quantitative analyses (Fig. 2). A Support Vector Machine (SVM) boundary computed over the entire dataset was used to evaluate separability of G≥4 tissue signature and healthy tissue (Table 2, basic separability). To assess whether FLIm data could effectively discriminate between healthy and PCa tissue, we evaluated SVM using average lifetimes from individual bands as well as the entire set of parameters obtained from the Laguerre deconvolution approach. A leave-one-patient-out approach was implemented to prevent data leakage. ROC-AUCs (Receiver Operating Characteristic Area Under the Curve) were computed across the entire dataset by first determining a probability for each class and generating an ROC curve using all patients. For the 3-class prediction framework, we generated ROC curves for each tissue class using a one-versus-rest approach. Specifically, we compared the predicted probability of each target class against the maximum probability from the remaining two classes, allowing for evaluation of classification performance across all tissue categories while accounting for the multi-class nature of the problem. The number of point measurements varies across patients, due to number and length of biopsy cores. A patient-level weighting based on the number of points was applied to the overall ROC-AUC computation to ensure that each patient is represented equally. Finally, to evaluate the effect of data leakage on classification performance, as observed in earlier publications^13^. We also evaluated classification performance using a 10-fold cross-validation where folds were randomly determined irrespective of patients.

**Table 2.** Classification Performance for Different Tissue Discrimination Approaches.

| Discrimination | Basic separability | 2-class prediction |  | 3-class prediction |  |
| --- | --- | --- | --- | --- | --- |
| Tissue class | PCa G4+<br>Benign | PCa (G3+G4+G5)<br>Benign | PCa (G4+G5)<br>PCa G3+Benign | PCa (G4+G5)<br>PCa G3<br>Benign | PCa (G4+G5)<br>PCa G3<br>Benign |
| Validation approach | No separate training/<br>validation sets | Leave-one-patient-out cross validation |  |  |  |
| Parameters | 470-nm LT<br>630-nm LT | 390-nm LT<br>470nm LT<br>540-nm LT<br>630-nm LT | 390-nm LT<br>470nm LT<br>540-nm LT<br>630-nm LT | 390-nm LT<br>470nm LT<br>540-nm LT<br>630-nm LT | LT and Laguerre<br>coefficients for all<br>spectral bands (n=52<br>parameters) |
| ROC-AUC | 0.950 | 0.835 | 0.887 | Benign: 0.833<br>Grade 3: 0.727<br>Grade 4+: 0.882 | Benign: 0.772<br>Grade 3: 0.711<br>Grade 4+: 0.789 |
| Sensitivity/<br>Specificity | 86.4%/ 90.1% | 84.4%/ 62.9% | 79.3%/85.0% | Benign: 68.0%/81.2%<br>Grade 3: 82.7%/55.2%<br>Grade 4+: 78.8%/80.5% | Benign: 67.2%/73.5%<br>Grade 3: 82.3%/50.9%<br>Grade 4+: 71.2%/71.2% |

## RESULTS

### Measurement Repeatability

The repeatability analysis demonstrated excellent consistency across sequential FLIm measurements. Quantitatively, only 3.3% of all scan points showed statistically significant differences between repeat measurements when evaluated at a 99% confidence threshold (Figure S2 B). This degree of consistency was maintained across all four spectral bands, underscoring the robustness of measurement stability. Furthermore, for all but a single isolated case (involving a single spectral band in one scan), the magnitude of variation remained within the inherent noise level of the system (See Figure S2 C). This high level of repeatability confirms that variations in FLIm signature during the brief measurement period are minimal and therefore unlikely to introduce confounding factors that would adversely affect tissue classification performance.

### Multivariate Analysis

Examination of the FLIm parameter space revealed distinct clustering patterns between tissue types. Most notably, benign prostatic tissue and Gleason Pattern 4 PCa demonstrated clear separation using only two parameters, using for example the 470-nm and 630-nm lifetime values (Figure 2A). This separation is particularly significant as it indicates that high-grade cancer can be distinguished using a minimal set of FLIm parameters. In contrast, Grade 3 cancer tissue demonstrated considerable overlap with both benign and Grade 4 signatures, suggesting intermediate biologic characteristics that are reflected by the FLIm measurements. When focusing specifically on Grade 4 patterns, no distinct differences were observed between cribriform and non-cribriform subtypes using our current parameter set. Quantitatively, applying a SVM classifier to the entire dataset based solely on these two parameters, yielded an ROC-AUC of 0.95 for distinguishing Grade 4 PCa from benign tissue (Table 2), demonstrating the strong diagnostic potential of this approach even with a simplified parameter set.

### Tissue Predictor Performance

Table 2 provides a comprehensive comparison of classification performance across multiple tissue discrimination scenarios and parameter combinations. Consistent with the multivariate analysis findings, the highest classification accuracy was achieved when discriminating Grade ≥4 PCa from benign and Grade 3 tissue (ROC-AUC: 0.887), while Grade 3 cancer showed more limited separability due to its intermediate FLIm signature characteristics. This performance hierarchy aligns with the biologic continuum from benign tissue to increasingly aggressive cancer phenotypes. Notably, we found that using just four average lifetime parameters (one from each spectral band) outperformed a more complex SVM model that utilizes the full set of 52 lifetimes and Laguerre expansion parameters. This finding suggests that the larger model may be prone to overfitting given the limited dataset, whereas the reduced parameter set offers more robust and generalizable classification. performance. On the other hand, classifiers with additional parameters, combined with a more expansive dataset, may help with discrimination of Grade 3 or Grade 4 cribriform vs. Grade 4 non-cribriform. When comparing validation methodologies, we observed that the conventional 10-fold cross-validation approach (which does not respect patient boundaries) produced an artificially inflated discrimination performance (ROC-AUC: 0.97), further validating our decision to implement the more rigorous leave-one-patient-out validation strategy to obtain realistic performance estimates.

## DISCUSSION

This work establishes FLIm as a promising approach for real-time discrimination of high-grade PCa, representing an important step toward integrating this technology into prostate biopsy devices for improved cancer detection during standard clinical procedures.

### Diagnostic Performance Implications

Multivariate analysis revealed consistent, diagnostically relevant contrast between benign and cancerous prostate tissue, with highest discrimination observed between Grade ≥4 PCa and benign tissue. The observed overlap between tissue type distributions likely stems partially from registration challenges between fresh specimens and histological sections, establishing a practical upper limit to achievable classification accuracy. Nevertheless, our classifier demonstrated robust performance in distinguishing PCa from benign tissue (ROC-AUC: 0.82) and particularly in identifying high-grade PCa (ROC-AUC: 0.88). The ability to specifically detect Grade ≥4 PCa has substantial clinical relevance, since more than minimal percentages of high-grade tumor usually render a patient ineligible for active surveillance. By providing real-time identification of high-grade cancer during the biopsy procedure, FLIm technology may improve targeting of clinically significant cancer, enhancing both diagnostic accuracy and treatment planning.

### Biological Basis of FLIm Contrast

The diagnostic potential of FLIm stems from its ability to capture simultaneously two fundamental biological changes in cancerous tissue. First, structural remodeling of the extracellular matrix occurs both within tumors and their surrounding microenvironment^18 19^, producing detectable changes in the 390-nm emission band through altered collagen and proteoglycan fluorescence signatures. Second, cancer cells undergo characteristic metabolic reprogramming through the Warburg effect—prioritizing glycolysis and lactate production even in oxygen-rich environments while reducing dependence on oxidative phosphorylation. This metabolic shift disrupts the equilibrium between free and protein-bound forms of NADH and FAD^20–22^, creating distinctive fluorescence lifetime patterns. Our data demonstrates this metabolic shift through the decreased fluorescence lifetime observed in the 470-nm band of tumor tissue, consistent with elevated levels of free NADH typical of glycolytic metabolism. Complementary metabolic information is captured in the 540-nm band through FAD fluorescence variations, while the 630-nm band detects changes in lipofuscin, an age-related pigment previously linked to PCa’s optical signature^13^. This multi-spectral approach enables comprehensive characterization of prostate tissue through simultaneous assessment of both structural and metabolic alterations associated with malignancy.

### Translational Relevance of Ex Vivo Findings

A major strength of our approach was acquiring measurements within seconds of biopsy collection. This was made possible by placing the instrumentation directly in the procedure room and implementing a rapid measurement protocol that preserved tissue integrity. This represents a methodological improvement over prior studies on prostatectomy specimens, where tissue was imaged an hour or more after blood supply termination— conditions likely to alter metabolic fluorescence signatures. The consistency of FLIm parameters observed in our repeat measurements (separated by up to 25 seconds) provides strong evidence that the optical signatures remain stable during the brief interval between specimen collection and measurement. This temporal stability suggests that the FLIm signatures we observed are likely representative of *in vivo* tissue characteristics.

### Methodological Improvements Over Prior Work

Several aspects of our study design have addressed important limitations identified in previous reports. Unlike earlier studies^13^ that analyzed predetermined regions of interest, our analysis incorporated point measurements along the entire length of biopsy cores, providing a more comprehensive assessment of tissue heterogeneity^13^. Furthermore, we implemented patient-level separation in our validation strategy, avoiding the methodological pitfall of traditional K-fold cross-validation approaches that, when applied at the point level, can lead to training and testing on highly correlated measurements from the same patient. This correlation between spatially adjacent measurements artificially inflates reported performance metrics and compromises model generalization. The “leave-one-patient-out” cross-validation approach employed in our analysis provides a more realistic representation of clinical performance, supported by consistent results obtained across multiple classification algorithms and parameter combinations.

### Study Limitations and Future Work

While our cohort of 20 patients was sufficient to demonstrate preliminary efficacy, validation on a larger and more diverse patient population will be necessary to establish definitive classification performance. Additionally, although our *ex vivo* measurements showed remarkable consistency with minimal temporal variation, direct confirmation through comparative *in vivo* studies will be essential to fully validate the translational potential of this approach. Future work should also explore integration of the FLIm technology directly into biopsy instrumentation to enable real-time tissue characterization during the procedure itself.

## CONCLUSION

FLIm demonstrates strong potential for real-time PCa detection during biopsy procedures. Our study demonstrates that FLIm can reliably differentiate clinically significant (Grade ≥4) PCa from benign tissue (ROC-AUC: 0.88) by detecting intrinsic metabolic and ECM composition differences. The single-fiber approach is well-suited for integration into existing biopsy instruments, enabling guided procedures that improve diagnostic yield. Although additional validation is required, these findings indicate that FLIm technology could significantly contribute to PCa diagnosis, inform treatment decisions, and ultimately improved patient outcomes.

## Data Availability

All data produced in the present study are available upon reasonable request to the authors

## Acknowledgement

The authors wish to acknowledge the support of the UC Davis Comprehensive Cancer Center Biostatistics Shared Resource, supported by the National Cancer Institute of the National Institutes of Health under award number NCI P30CA093373. The content is solely the responsibility of the authors and does not necessarily represent the official views of the National Institutes of Health.

## Supplementary material

**Figure S1.**
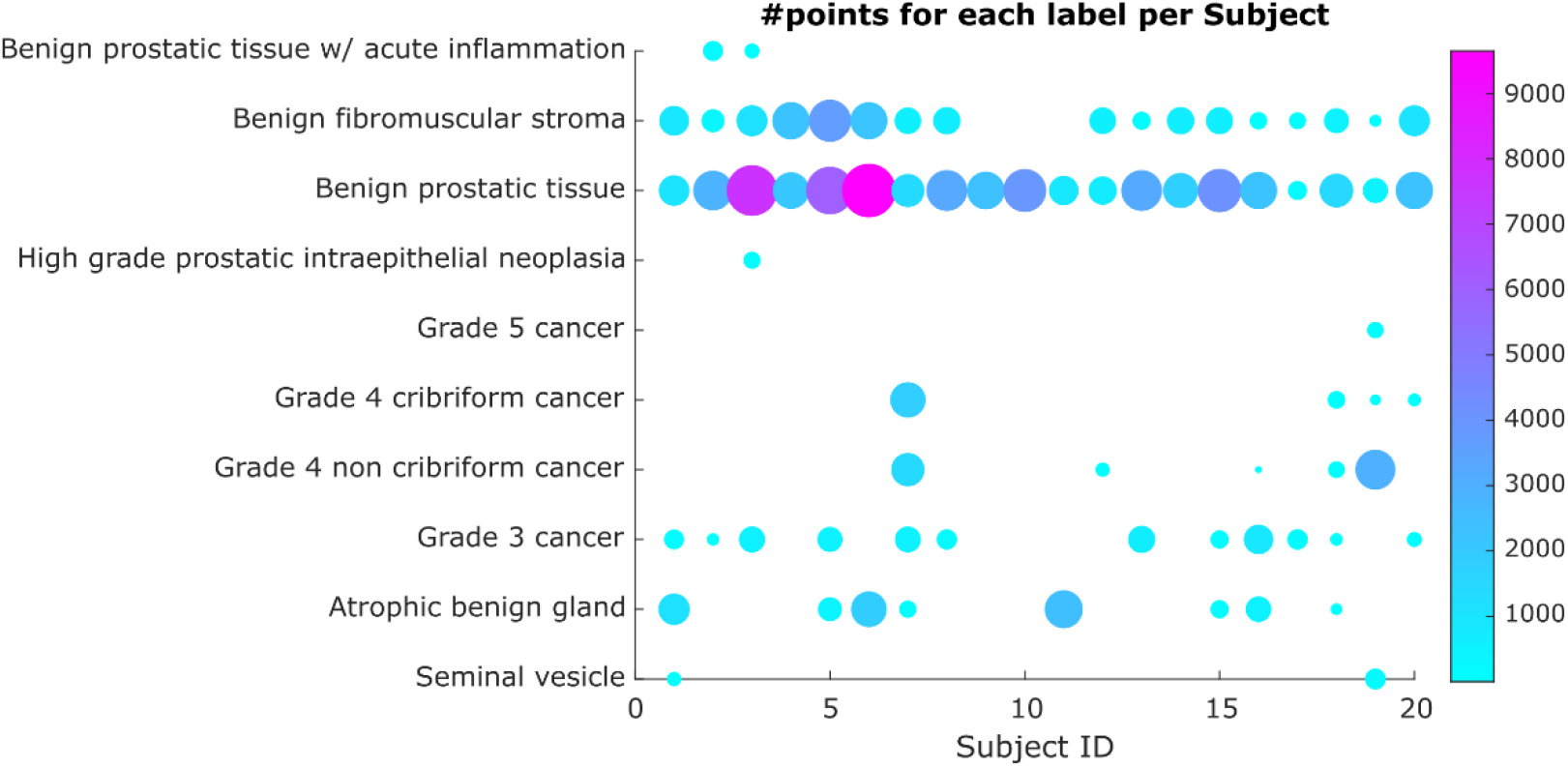
Representation of study database. Each dot represents the number of points for each label and each patient enrolled in the study.

**Figure S2.**
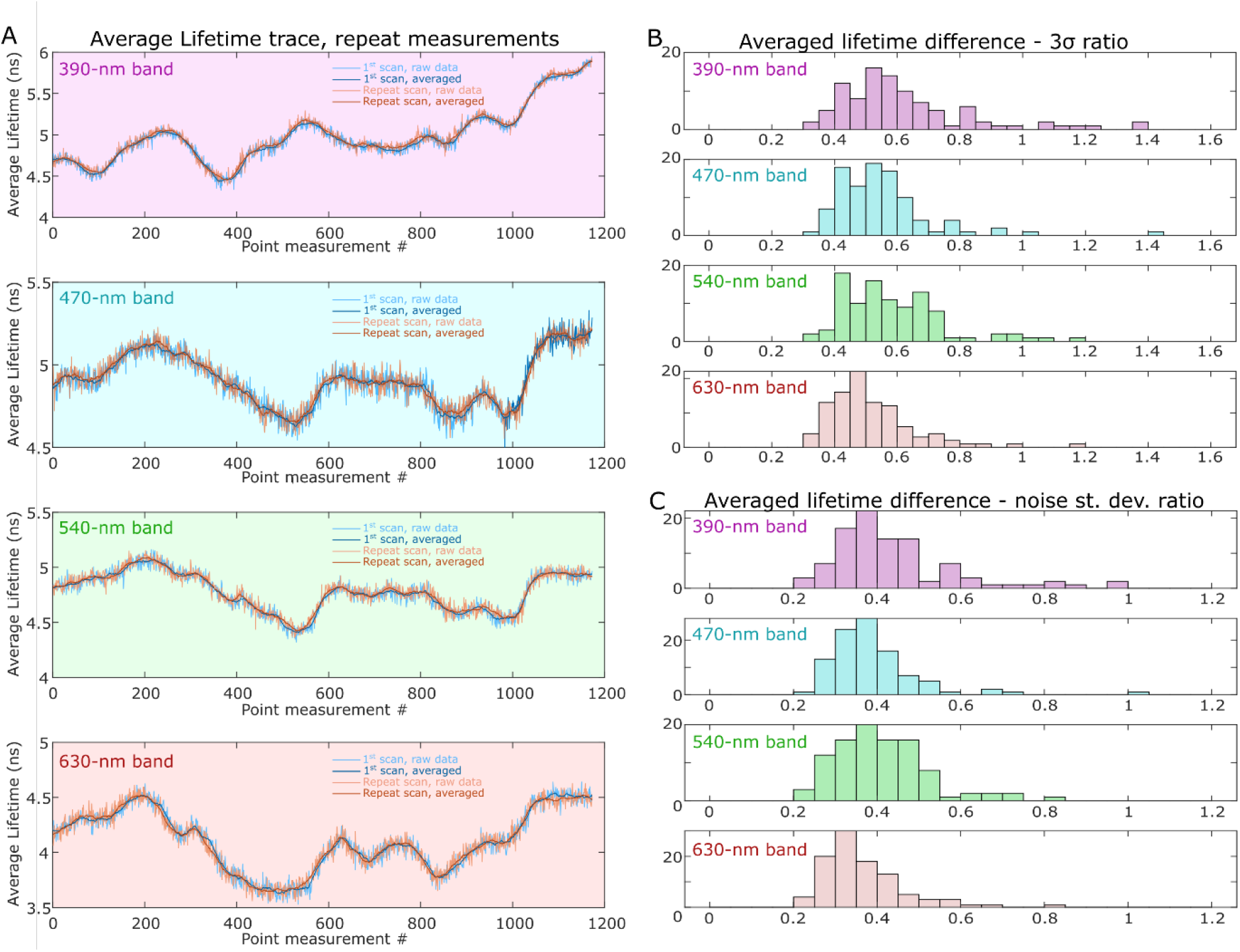
Measurement repeatability evaluation. Raw/smooth traces of average lifetime for the four spectral bands of the instrument (A). Ratio of mean absolute error and 3 standard deviations of the estimated noise in the smooth traces. Values above 1 show that the difference is unlikely to be due to measurement noise alone with a confidence interval of .99 (B). Ratio of mean absolute error and standard deviations of the estimated noise in the original traces demonstrating that differences in repeat scans are well below the measurement noise (c)

